# Risk factors associated with cardiovascular disease in Mexican people with Systemic Lupus Erythematosus

**DOI:** 10.1101/2025.06.19.25329956

**Authors:** Ana Laura Hernández Ledesma, Sofía Fernanda Hernández-Rodríguez, Ana Sofía Del Angel Zambrano, Grecia Sevilla-Parra, Angélica Peña-Ayala, Lizbet Tinajero-Nieto, Estefania Torres-Valdez, David Gustavo García-Gutiérrez, Adriana Jheny Rodríguez-Méndez, Jesús Sepúlveda-Delgado, Gilberto M. Vázquez-Mejía, Deshiré Alpizar-Rodríguez, Alejandra Medina-Rivera, Domingo Martínez

**Affiliations:** Laboratorio Internacional de Investigación sobre el Genoma Humano, Universidad Nacional Autónoma de México, Querétaro, Querétaro, México; Licenciatura en Medicina, Universidad del Valle de México, Querétaro, Querétaro, México; Licenciatura en Médico Cirujano, Universidad Anáhuac, Querétaro, Querétaro, México; Licenciatura en Neurociencias, Escuela Nacional de Estudios Superiores Unidad Juriquilla, Universidad Nacional Autónoma de México, Querétaro, Querétaro, México; Hospital General Regional No. 1, Instituto Mexicano del Seguro Social, Querétaro, Santiago de Querétaro, México; Instituto Nacional de Rehabilitación “Luis Guillermo Ibarra Ibarra”, Ciudad de México, México; Hospital General Regional No. 2, Instituto Mexicano del Seguro Social, El Marqués, México; Facultad de Química, Universidad Autónoma de Querétaro, Querétaro, México; Centro de Investigación Biomédica Avanzada, Facultad de Medicina, Universidad Autónoma de Querétaro, Querétaro, México; Research Division, Servicios de Salud IMSS BIENESTAR, Hospital Regional de Alta Especialidad, Ciudad Salud, Chiapas, México; Nephrology, private practice at Clínica Vázquez, Guanajuato, México

**Keywords:** Systemic lupus erythematosus, cardiovascular disease risk factors, cardiovascular disease risk factors associated with lupus, hypertension, lupus nephritis, prediction model

## Abstract

**Background:** Traditional cardiovascular risk factors are more frequent among people with Systemic Lupus Erythematosus (SLE); however, they cannot fully explain the increased risk for cardiovascular diseases among this population. Lupus-associated factors have been proposed to contribute to cardiovascular disease risk. For instance, hypertension is a major risk factor in lupus, and it is associated with renal outcome, including nephritis, a severe complication of lupus. This study aims to evaluate how traditional and lupus-associated risk factors contribute to cardiovascular disease in Mexican people with SLE.

**Methods:** A clinical interview, physical examination, and blood sampling were conducted in a Clinical cohort to explore, by two independent regression models, how traditional and lupus-associated factors contribute to cardiovascular disease risk using the Framingham risk score. Then, in a Registry cohort, with data from the Mexican Lupus Registry, we implemented two independent Bayesian network models to predict nephritis and hypertension, combining traditional and lupus-associated factors.

**Results:** Twenty-eight women were recruited, through medical consultation, in our clinical cohort, with a median age of 43.5 (14.25) years. Twelve (42.9%) reported previous cardiovascular conditions; seven (25%) had hypertension. According to the Framingham risk score, three (10.7%) showed low risk, eighteen (64.3%) mild risk, and seven (25%) high risk. Among traditional risk factors, one (3.6%) presented hypercholesterolemia (>200 mg/dL), and two (7.1%) high systolic pressure (>130 mmHg). Regarding lupus-associated factors, the majority use antimalarials (71.4%), two (7.1%) reported no activity of the disease, and seven (25%) had nephritis. In the regressions among traditional factors, systolic pressure and triglycerides showed significant effect; whereas for lupus-associated factors age at diagnosis, years with lupus, socioeconomic level, corticoids, and antimalarials were significant. Regarding Bayesian networks, using our Registry cohort with 2914 cases; treatment, diagnosis delay, years with lupus, damage accrual (SLICC), disease activity (SLEDAI), autoantibody test, ancestry, and relatives with lupus, showed causality over hypertension and nephritis. The nephritis network correctly identified 90% of the nephritis cases, whereas the hypertension network correctly identified 94% of the hypertension cases.

**Conclusion:** Additional to traditional cardiovascular risk factors, lupus-associated factors should be considered to improve the estimation of the cardiovascular risk score in the Mexican lupus population.

## 1 Introduction

Systemic lupus erythematosus (SLE) is a chronic inflammatory disease of multifactorial etiology, with incidence and prevalence that vary between countries (1). It can manifest at any age, but is more common in people aged between 15 and 60 years old, with a 9:1 ratio between women and men (1,2). Its clinical manifestations include cutaneous, joint, hematological, neuropsychiatric, renal, gastrointestinal, pulmonary, and cardiovascular symptoms (1,3,4). It is characterized by chronic inflammation and immune system dysfunction, which can contribute to vascular damage, atherosclerosis, and thrombosis. These complications significantly increase the risk of cardiovascular diseases (CVDs) and strokes, contributing to higher mortality and morbidity rates among people with SLE (5).

Compared to the general population, individuals with SLE have a two to ten times higher risk of experiencing cardiovascular events, which are associated with nearly a third of deaths among patients with over 10 years of diagnosis (5,6). Previous studies have reported that individuals with SLE have twice the risk of developing nonfatal cardiovascular events, with a hazard ratio for myocardial infarction ranging from 2.6 to 5.1, whereas for stroke, it ranges from 2.1 to 3.3. Annual rates for myocardial infarction, strokes, and atherosclerotic plaque formation are also increased among young individuals with SLE, compared to the general population (7,8). The risk factors for cardiovascular disease in SLE patients encompass both traditional elements, such as hypertension, diabetes mellitus, dyslipidemia, and smoking, as well as disease-specific factors like disease activity and the use of corticosteroids and immunosuppressants (7). For instance, CVDs are displacing infections as the leading cause of mortality among people with SLE, primarily due to the use of corticosteroids and immunosuppressants (8).

The mechanisms involved in the increased risk for atherosclerosis and CVDs in people with SLE have not been fully established, yet both cardiovascular risk factors and SLE-associated factors have been proposed to contribute (8). Cardiovascular risk factors (CRFs) are defined as biological characteristics or lifestyles that increase the probability of suffering or dying from cardiovascular diseases; those proven to be associated with CVDs are considered traditional factors, such as hypertension, dyslipidemia, obesity, diabetes, smoking, age, sex, and family history (9,10). In contrast, non-traditional factors include those identified in studies of cardiovascular diseases, but their contribution has not been fully determined, such as markers of chronic inflammation, physical activity, alcoholism, eating habits, gut microbiota, and psychosocial factors (9,10).

Among people with SLE, it has been reported that traditional CRFs are more frequent, along with higher cardiovascular risk scores, when compared with healthy controls (7,8). Furthermore, differences in levels of CRFs such as very low-density lipoprotein cholesterol (VLDL-C), triglycerides and high-density lipoprotein cholesterol (HDL-C) have also been observed between people with disease activity and those in remission (8). Other factors associated with higher CVDs risk among people with SLE include body mass index, familial history of cardiac disease, hypertension, sedentary lifestyle, premature menopause, diabetes mellitus/insulin resistance, and tobacco use (7,8). Although CRFs have been reported to contribute to CVDs, they cannot fully explain the increased risk among people with SLE. Therefore, factors associated with SLE diagnosis and management have been proposed as contributing to CVDs within this population (6–8). Among the factors contributing to CVDs in SLE, hypertension has been considered a major risk factor (11), and it has been associated with nephritis, another frequent and severe SLE-associated manifestation (12,13). Nephritis and hypertension highly increase morbidity and mortality in SLE (14–16).

Early identification of the susceptibility to develop CVDs has become essential for prevention and decreasing the potential effects of its appearance, which has led to the development of studies and scales to evaluate CRFs (11). The Framingham risk score (FRS) is one of the most used prediction tools, designed to help estimate the risk of having a cardiovascular event by considering CRFs such as age, gender, total cholesterol, systolic blood pressure, high-density lipoprotein (HDL-C) cholesterol, and smoking habit (17).

As previously described, FRS may have limitations for clinical populations, since it does not include key factors that have been shown to contribute to CVDs in people with SLE, such as the use of glucocorticoids, disease activity (e.g., SLEDAI), organ damage (especially renal), or the presence of autoantibodies(7,8,18). The exact role of CRFs and SLE-associated factors in the etiology of CVDs in people with SLE remains unclear. To know their contribution, will allow us to identify potential markers, leading to design new strategies that favor the early detection of CVDs risk among people with SLE. To evaluate how cardiovascular and SLE-related factors contribute to disease risk in Mexican individuals with SLE, we analyzed two complementary cohorts. The first, a **Clinical Cohort** (n=28), consisted of in-person participants who underwent clinical evaluation, blood sampling, and cardiovascular risk assessment using the Framingham Risk Score (19). The second, a **Registry Cohort**, includes over 2,900 individuals recruited virtually via the LupusRGMX platform. This cohort was used to construct Bayesian network models to predict nephritis and hypertension, integrating a broad set of clinical, demographic, and disease-specific variables.

## 2 Materials and methods

### 2.1 Study design and population

This is a cross-sectional study nested in a cohort. We evaluated the frequency of CRFs and SLE-associated factors and their contribution to the development of CVDs in a population of Mexican people with SLE. The project was approved by the Research Ethics Committee of the Institute of Neurobiology at Universidad Nacional Autonoma de Mexico (UNAM). All the data was anonymized and stored at the National Laboratory of Advanced Scientific Visualization from UNAM. The participants provided digitally or on-site signed informed consent and received a copy of the privacy statement, in accordance with the Federal Law on the Protection of Personal Data Held by Private Parties.

Volunteers were recruited by rheumatologists affiliated to LupusRGMX (Clinical cohort), and through the LupusRGMX platform (Registry cohort) (20), between May 2021 and September 2024. All volunteers met the criteria of the American College of Rheumatology and the Systemic Lupus Erythematosus International Collaborating Clinics criteria (ACR/SLICC). Only participants aged over 18 years old, and with a confirmed diagnosis of SLE, were included. Participants with incomplete records were excluded from the analyses. Details about LupusRGMX had been published elsewhere (21,22).

### 2.2 Framingham Risk Score estimation

The Clinical cohort of women was recruited by rheumatologists affiliated with LupusRGMX. A clinical interview and a physical examination were conducted to explore risk factors, including previous diabetes mellitus diagnosis, use of antihypertensives, lifestyle (which comprises use of alcohol, cigarettes, eating behavior and physical activity), as well as to assess of vital signs (heart rate, respiratory rate and blood pressure), with emphasis on blood pressure. Hypertension was determined when systolic and diastolic pressure were over 130 and 90 mmHg, respectively; in accordance with the American Heart Association (23). Weight and height were also measured to calculate the body mass index (BMI); individuals were considered overweight when their BMI was higher than 25.

Blood samples were also collected after an 8-hour fast. A basic metabolic panel test was performed using the A15 Analyzer (BioSystems); this panel includes total cholesterol, triglycerides (TG), high-density lipoprotein (HDL-C) and low-density lipoprotein cholesterol (LDL-C). Reference values for the Mexican population were considered for the interpretation of each biomarker (22).

The estimation of cardiovascular risk was determined using the modified Framingham score provided by the Instituto Mexicano del Seguro Social (IMSS), as it is a standard tool for estimating the risk of CVDs in the Mexican population (24,25). This calculator considers age, sex, total cholesterol and HDL levels, smoking habit, the presence, or absence of diabetes mellitus, systolic blood pressure level, and the use of antihypertensive medications. With these variables, the Framingham score calculator estimates a person’s risk percentage of experiencing a cardiovascular event within the next 10 years, and classifies them as low (<1%), moderate (1-5%), or high (>5%) risk.

### 2.3 Contribution of CRFs and SLE-associated factors to Framingham Risk Score estimation

Once we estimated the risk of CVDs in our Clinical Cohort, we performed and adjusted two independent linear regression models. The first model aimed at predicting CVDs risk from CRFs; whereas the second model aimed at predicting CVDs risk from SLE-associated factors. For both cases, we ran a typical regression, then we ran a step backward regression to select the best predictors. Finally, we checked consistency and collinearity among predicting variables for each model; if needed, we removed closely related variables. Moreover, we verified consistency by a permutation test, in such inconsistent variables. Regarding disease activity, people over 12 SLEDAI points were classified as high or severe disease activity, whereas the rest of people were classified as low or moderate disease activity (26). If inconsistencies were statistically significant, it suggested that such inconsistencies were unlikely to have occurred by chance. The complete analysis code is avialibale through GitHub https://github.com/NeuroGenomicsMX/Cardiovascular_Risk_Factors_Lupus_Mex.

In the first model, we set the following CRFs as predictor variables: total cholesterol, triglycerides, HDL-C, LDL-C, tobacco use, alcohol use, systolic and diastolic pressure, heart rate, and respiratory rate. Due to the sample size, we selected less than ten variables. In the second model, we set the following SLE-associated factors as predicting variables: hypertension, disease duration, socioeconomic level, lupus nephritis, antihypertensive medications consumption, antimalarials consumption, corticoids consumption, and disease activity (SLEDAI).

### 2.4 Predictive model for nephritis and hypertension

With the data from our Registry Cohort (21) we developed predictive models for CVDs risk components, specifically, nephritis and hypertension, for people with SLE. We selected these as target variables to be predicted, as hypertension is one of the most prevalent risk factors for CVDs (11,20) and it is strongly linked to nephritis (13,17); both of them highly increase morbidity and mortality in SLE (14–16). Thus, the second cohort was assembled using the Mexican Lupus Registry, with a total of 2,914 participants.

For each target variable, nephritis and hypertension, we implemented a Bayesian network model (27), i.e., one independent model to predict lupus nephritis, and another independent model to predict hypertension. We used the bnlearn package (28) to implement the Bayesian network models. All the analyses were developed and implemented in R language, version 4.4.3, and are available on https://github.com/NeuroGenomicsMX/Cardiovascular_Risk_Factors_Lupus_Mex. All data is available upon request through the MexOMICS Consortium (https://redcap.link/nqsxtj8n). Bayesian networks were chosen because they allow to combine data-driven causality (learnt from the network’s structure and parameters) with expert knowledge (29,30) from rheumatologists and nephrologists; in addition, they work well with noisy and incomplete data (31–33), and they are easy to interpret because of the fully connected probabilistic graph (30,34).

First, from the completed data, we integrated a dataset for lupus nephritis and another independent dataset for hypertension, selecting the most important variables previously identified. After removing incomplete cases, we kept 1256 cases for the nephritis data set, and 911 cases for the hypertension data set. Since nephritis and hypertension showed unbalanced classes, we balanced them by the synthetic minority over-sampling technique, SMOTE (35–37). Therefore, we got a balanced nephritis data set of 486 cases and a balanced hypertension data set of 606 cases. Each data set was split, 70% for training models and 30% for validation.

In both cases, as predictive or explanatory variables, we considered 529 variables, including CRFs and SLE-associated factors. Then, we ran the Wrapper Algorithm for all relevant features’ selection, using the Boruta package (38,39). Then, we ran and optimized a random forest model to identify other potential predictive variables for the nephritis Bayesian model and other potential predictive variables for the hypertension Bayesian model.

Regarding random forest models, we trained both of them running a search grid with the following hyperparameters: number of trees = 10 times number of predictive variables; sampling with and without replacement; resampling fraction: {0.4, 0.5, 0.6, 0.7, 0.8}; minimum node size: {1, 3, 5, 10, 20, 30, 40, 50}; mtry: {0.05, 0.15, 0.25, 0.33, 0.4, 0.6, 0.7, 0.8, 1} times number of predictive variables. Once we obtained the best parameters, we reran each model with these optimal parameters and 5000 trees to calculate the importance of the variables using both the permutation approach and the impurity approach. Thus, based on such importance, we identified which variables could be inputs for the nephritis Bayesian model and which ones could be inputs for the hypertension Bayesian model. We integrated such variables with those results derived from the Boruta package.

In the next step, to learn a provisional structure of both networks, using the important variables previously identified, we ran the Grow-Shrink algorithm, a constraint-based approach, and the Reconstruction of Accurate Cellular Network algorithm, which is a mutual information-based algorithm. Such provisional network structures were presented to rheumatologists and nephrologists for validation. Therefore, based on their clinical experience, as well as on what they knew was reported in scientific literature, they kept, removed, or added variables and relations (i.e., conditional dependencies, also called causalities). Once they reached an agreement for the nephritis network structure and for the hypertension network structure, we fitted the models to predict nephritis and hypertension, with such network structures, respectively. Finally, we evaluated model performance for both networks in the validation dataset, respectively. Performance metrics included accuracy, precision, recall, and an F1 score.

## 3 Results

### 3.1 CRFs and SLE factors are both informative for CVDs risk estimation

A total of twenty-eight women were recruited as volunteers in our Clinical cohort, with a median age of 43.5 (14.25) years; the median age at diagnosis was 31 (20.75) years. Demographic, lifestyle, and clinical characteristics of the volunteers are summarized and described in Table 1.

**Table 1.** Sociodemographic, clinical, lifestyle, and cardiovascular disease risk factors in the Clinical cohort (n=28).

| *Variables* | *Total*<br>*(n=28)* | *Low*<br>*CVDR (n=3)* | *Mild CVDR*<br>*(n=18)* | *High CVDR*<br>*(n=7)* |
| --- | --- | --- | --- | --- |
| ***Framingham scale*** |  |  |  |  |
| Cardiovascular disease risk, median (IQR) | 2.4 (2.75) | 0.0 (0.00) | 2.2 (1.3) | 6.3 (2.8) |
| ***Sociodemographic*** |  |  |  |  |
| Age, median (IQR) | 43.5 (14.25) | 29.0 (3.50) | 42.5 (9.25) | 63.0 (11.50) |
| Age of diagnosis, median (IQR) | 31 (20.75) | 19 (2.50) | 30 (11.75) | 51 (14.00) |
| Socioeconomic level (n,%) |  |  |  |  |
| C- (middle) | 5 (17.9%) | 1 (33.3%) | 4 (22.2%) | 0 (0%) |
| D+ (middle) | 8 (28.6%) | 0 (0%) | 6 (33.3%) | 1 (28.6%) |
| D (low) | 15 (53.6%) | 2 (66.7%) | 8 (44.5%) | 5 (71.4%) |
| ***Cardiovascular disease*** |  |  |  |  |
| Hypertension (n,%) | 7 (25%) | 0 (0%) | 4 (22.2%) | 3 (42.9%) |
| Heart failure (n,%) | 1 (3.6%) | 0 (0%) | 0 (0%) | 1 (14.3%) |
| Venous insufficiency (n,%) | 8 (28.6%) | 0 (0%) | 3 (16.7%) | 5 (71.4%) |
| Deep vein thrombosis (n,%) | 3 (10.7%) | 0 (0%) | 2 (11.1%) | 1 (14.3%) |
| ***Clinical*** |  |  |  |  |
| Disease duration, median (IQR) | 8 (9.25) | 9 (1.5) | 8 (14.75) | 5 (7.50) |
| Disease activity |  |  |  |  |
| No activity (n,%) | 2 (7.1%) | 0 (0%) | 2 (11.1%) | 0 (0%) |
| Low activity (n,%) | 2 (7.1%) | 0 (0%) | 1 (5.6%) | 1 (14.3%) |
| Moderate activity (n,%) | 2 (7.1%) | 0 (0%) | 2 (11.1%) | 0 (0%) |
| High activity (n,%) | 11 (39.3%) | 2 (66.7%) | 5 (27.8%) | 4 (57.1%) |
| Very high activity (n,%) | 11 (39.3%) | 1 (33.3%) | 8 (44.4%) | 2 (28.6%) |
| ***Lipid profile*** |  |  |  |  |
| HDL-C, median (IQR) | 41.5 (15.00) | 41.0 (11.5) | 41.5 (12.75) | 42.0 (14.00) |
| Triglycerides, median (IQR) | 74.0 (35.0) | 58.0 (9.0) | 78.5 (34.5) | 73.0 (42.5) |
| Total cholesterol, median (IQR) | 119.5 (31) | 111.0 (10) | 122.0 (28) | 115.0 (55) |
| LDL-C, median (IQR) | 60 (24.50) | 51 (7.00) | 66 (22.25) | 47 (32.00) |
| Familial SLE in FDR (n,%) | 6 (21.4%) | 0 (0%) | 4 (22.2%) | 2 (28.6%) |
| Familial SLE in SDR (n,%) | 2 (7.1%) | 0 (0%) | 2 (11.1%) | 0 (0%) |
| Familial HAS in FDR (n,%) | 17 (60.7%) | 3 (100%) | 10 (55.6%) | 5 (71.4%) |
| Familial HAS in SDR (n,%) | 12 (42.9%) | 3 (100%) | 8 (44.4%) | 2 (28.6%) |
| Diabetes (n,%) | 1 (3.6%) | 0 (0%) | 0 (0%) | 1 (14.3%) |
| Systolic pressure, median (IQR) | 110 (18.50) | 109 (10.50) | 110 (17.75) | 113 (19.00) |
| Diastolic pressure, median (IQR) | 78.5 (12.0) | 74.0 (10.5) | 79.0 (10.0) | 79.0 (13.5) |
| Body mass index, median (IQR) | 25.7 (9.450) | 24.2 (8.450) | 25.0 (5.175) | 33.2 (5.700) |
| Nephritis (n,%) | 7 (25%) | 1 (33.3%) | 3 (16.7%) | 2 (28.6%) |
| **Lifestyle** |  |  |  |  |
| Smoking (n,%) | 4 (14.3%) | 0 (0%) | 1 (5.6%) | 4 (57.1%) |
| Alcohol use (n,%) | 9 (32.1%) | 1 (33.3%) | 6 (33.3%) | 5 (71.4%) |
| Physical activity |  |  |  |  |
| Low activity (n,%) | 7 (25%) | 1 (33.3%) | 5 (27.8%) | 1 (14.3%) |
| Moderate activity (n,%) | 8 (28.6%) | 1 (33.3%) | 4 (22.2%) | 3 (42.9%) |
| High activity (n,%) | 10 (35.7%) | 0 (0%) | 8 (44.4%) | 2 (28.6%) |
| Unknown (n,%) | 3 (10.7%) | 1 (33.3%) | 1 (5.6%) | 1 (14.3%) |
| **Treatments** |  |  |  |  |
| Corticoids (n,%) | 18 (64.3%) | 1 (33.3%) | 11 (61.1%) | 6 (85.7%) |
| Antimalarials (n,%) | 20 (71.4%) | 3 (100%) | 13 (72.2%) | 4 (57.1%) |
| Antihypertensives (n,%) | 4 (14.3%) | 0 (0%) | 3 (16.7%) | 1 (14.3%) |

According to the FRS, only three (10.7%) of our volunteers were considered with low CVDs risk (FRS <1); whereas eighteen (64.3%) were classified with mild risk (FRS= 1-5), and seven (25%) with high CVDs risk (FRS>5). The median cardiovascular disease risk score, according to the FRS, was 2.4 (2.75) points, with a notable increase to 6.3 (2.8) points in the high CVDs group. The median heart age of the volunteers was calculated at 39 (18.75), while the real median age of the volunteers was 43.5 (14.25), the range of real age was lower than the estimated heart age.

Twelve (42.9%) of the volunteers reported a previous CVDs: seven (25%) had hypertension, one (3.6%) heart failure, eight (28.6%) venous insufficiency and three (10.7%) deep vein thrombosis. When analyzing the frequency of these CVDs among the three FRS groups, we observed that none of the low CVDs risk had a previous CVDs, in the mild CVDs risk group, one volunteer had diagnosis for hypertension (3.6%), two for deep vein thrombosis (7.1%), and three had both hypertension and venous insufficiency diagnosis (10.7%). In the high CVDs risk group, one volunteer had hypertension (3.6%), two had venous insufficiency (7.1%), two had hypertension and venous insufficiency (7.1%), and one had venous insufficiency, heart failure, and deep vein thrombosis (3.6%). Volunteers with hypertension showed a FRS median of 3.9 (3.65), in contrast with those without hypertension of 2 (1.3); for people with venous insufficiency, the median FRS was 5.8 (4.85), contrasting with those without such diagnosis of 1.7 (1.075).

Regarding the prevalence of CRFs we assessed the serum lipid profile of the volunteers; only one (3.6%) presented hypercholesterolemia (>200 mg/dL), three (10.7%) had hypertriglyceridemia (>150 mg/dL), twelve (43%) had low levels of HDL (<40 mg/dL) and two (7.1%) had high levels of LDL (>100 mg/dL). For blood pressure, only two volunteers (7.1%) presented high systolic pressure (>130 mmHg) and only one (3.6%) was on the upper limit for diastolic pressure (90 mmHg). We also identified that only one (3.6%) of the volunteers had a diabetes diagnosis, four (16%) reported smoking and seven (28%) reported consuming alcohol daily. When asked about their regular physical activity, seven (25%) of the volunteers reported a low activity, eight (28.6%) moderate and ten (35.7%) a high activity. According to the estimation of BMI, five of the volunteers were considered to have an overweight (≥25), and ten with obesity (≥30). Concerning family history of hypertension, it was observed in 60.7% of first-degree relatives and 42.9% of second-degree relatives.

Concerning the SLE-associated factors, we estimated a median disease duration of 8 (9.25) years, with a median age at diagnosis of 31 (20.75). Only seven (25%) had been diagnosed with nephritis. Antimalarials are the most frequent treatment among our volunteers (71.4%), followed by corticoids (64.3%); four of the volunteers reported to use antihypertensives regularly. Familiar history of SLE was reported on 21.4% of the volunteers.

Once the risk of CVDs and the heart age were estimated for our Clinical cohort using Framingham Score, we applied two regression models to predict, independently, the risk of CVDs using the same cohort (n=28). Specifically, regression model-1 was used to understand how CRFs explain CVDs risk (Table 1), whereas, regression model-2 shows how SLE-associated factors explain CVDs risk (Table 3).

Regarding CRFs as explanatory variables of CVDs risk in regression model-1, we initially included total cholesterol, triglycerides, high-density lipoprotein, low-density lipoprotein, tobacco use, alcohol use, systolic and diastolic pressure, heart rate, and respiratory rate. After the step backward regression, we found a significant model (p-value: 0.00182) with an adjusted R-squared of 0.5173, keeping total cholesterol, triglycerides, systolic and diastolic pressure, respiratory rate, and heart rate in the model (Table 2). It is worth mentioning that low-density lipoprotein was excluded because of collinearity with total cholesterol. Then, only tobacco use, systolic pressure, total cholesterol, and triglycerides showed a significant effect on CVDs risk. However, since tobacco use and total cholesterol showed a protective effect (see Table 2), which is counterintuitive and contrary to the literature, we explored in detail both variables.

**Table 2.** Regression model-1 results, CRFs explaining CVDs risk in the clinical cohort (n=28).

| *Predicting Variables* | *$\beta$ Estimate* | *Std. Error* | *P-value* |
| --- | --- | --- | --- |
| (Intercept) | 18.37538 | 6.07395 | 0.00668 |
| Tobacco use | -7.56928 | 1.40795 | 2.91e-05 |
| Systolic pressure | 0.12301 | 0.03809 | 0.00420 |
| Total cholesterol | -0.05694 | 0.02028 | 0.01087 |
| Triglycerides | 0.02756 | 0.01201 | 0.03262 |
| Diastolic pressure | -0.07573 | 0.04804 | 0.13064 |
| Respiratory rate | 0.03372 | 0.02296 | 0.15742 |
| Heart rate | -0.06584 | 0.04761 | 0.18195 |

*Residual standard error: 1.973 on 20 degrees of freedom* *Multiple R-squared: 0.6424, Adjusted R-squared: 0.5173* *F-statistic: 5.134 on 7 and 20 DF, p-value: 0.00182*

Regarding cholesterol levels, despite their elevated CVDs risk scores, we found only one person with hypercholesterolemia; the rest of them had normal levels. Regarding the protective effect of tobacco use in the linear model, we performed a permutation test, comparing the average CVDs risk between smoking and non-smoking participants. The differences between groups resulted in significant, i.e., smoking participants had a CVDs risk greater than non-smoking participants (see Figure 1) by 4.1125 points (on average, one-tail test, p-value: 0.008645), which contradicts regression model-1 (see Table 2).

**Figure 1.**
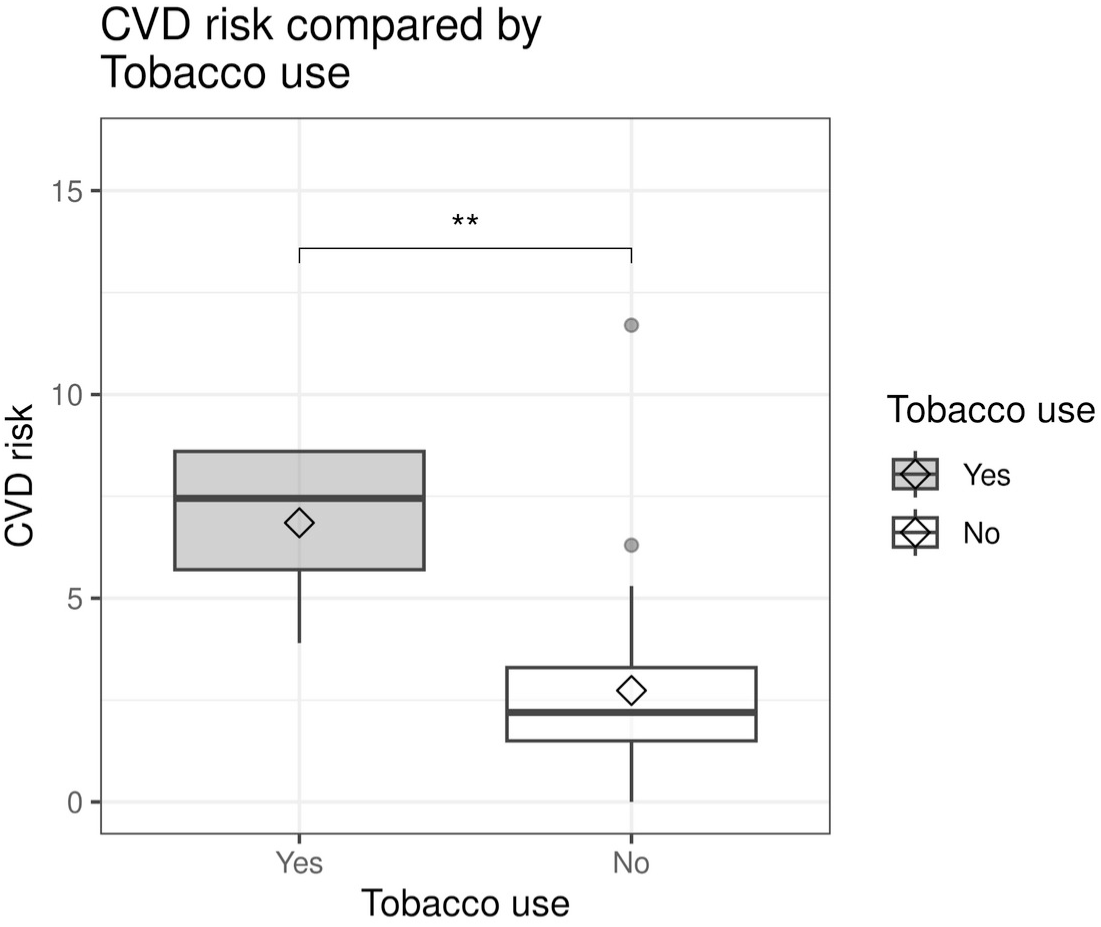
CVDs risk comparison for tobacco use in the Clinical cohort (n=28). For our Clinical cohort, smoking participants had a CVDs risk greater than non-smoking participants by 4.1125 points (on average, one-tail test, p-value: 0.008645), which highlights inconsistencies when only traditional CRFs are used to estimate CVDs risk in Mexican people with SLE.

We assessed SLE-associated factors as explanatory variables of CVDs risk in regression model-2, we included age at diagnosis, years with lupus, socioeconomic level, antimalarials consumption, corticoids consumption, disease activity, and nephritis. After the step backward regression, we found a significant model (p-value: 2.173e^-07^) with an adjusted R-squared of 0.7955, keeping all variables except nephritis, which was removed from the model. In this case, only Disease activity showed a protective effect, which is contrary to the literature (see Table 3).

**Table 3.** Regression model-2 results, SLE-associated factors explaining CVDs risk in the Clinical cohort (n=28).

| *Predicting Variables* | *$\beta$ Estimate* | *Std. Error* | *P-value* |
| --- | --- | --- | --- |
| (Intercept) | 1.585901 | 1.593948 | 0.33109 |
| Age at diagnosis | 0.145734 | 0.025283 | 1.01e-05 |
| Antimalarials consumption | -2.060494 | 0.571934 | 0.00167 |
| Years with lupus | 0.082357 | 0.027911 | 0.00763 |
| Socioeconomic level | -0.025390 | 0.009562 | 0.01481 |
| Corticoids consumption | 1.625151 | 0.615210 | 0.01526 |
| Disease activity | -0.044538 | 0.017951 | 0.02164 |
| *Residual standard error: 1.284 on 21 degrees of freedom* |  |  |  |
| *Multiple R-squared: 0.8409, Adjusted R-squared: 0.7955* |  |  |  |
| *F-statistic: 18.5 on 6 and 21 DF, p-value: $2.173e^{-07}$* |  |  |  |

However, this counterintuitive effect was verified by a permutation test, comparing mean CVDs risk between participants with high disease activity vs. participants with low disease activity, people over 12 SLEDAI points were classified as high disease activity. The differences between groups resulted no significant (see Figure 2), i.e., there is no CVDs risk significant differences between groups (two-sided test, p-value: 0.8513), which suggests that, disease activity effect in regression model-2 (see Table 3) is just a marginal effect that would be expected to disappear when increasing sample size.

**Figure 2.**
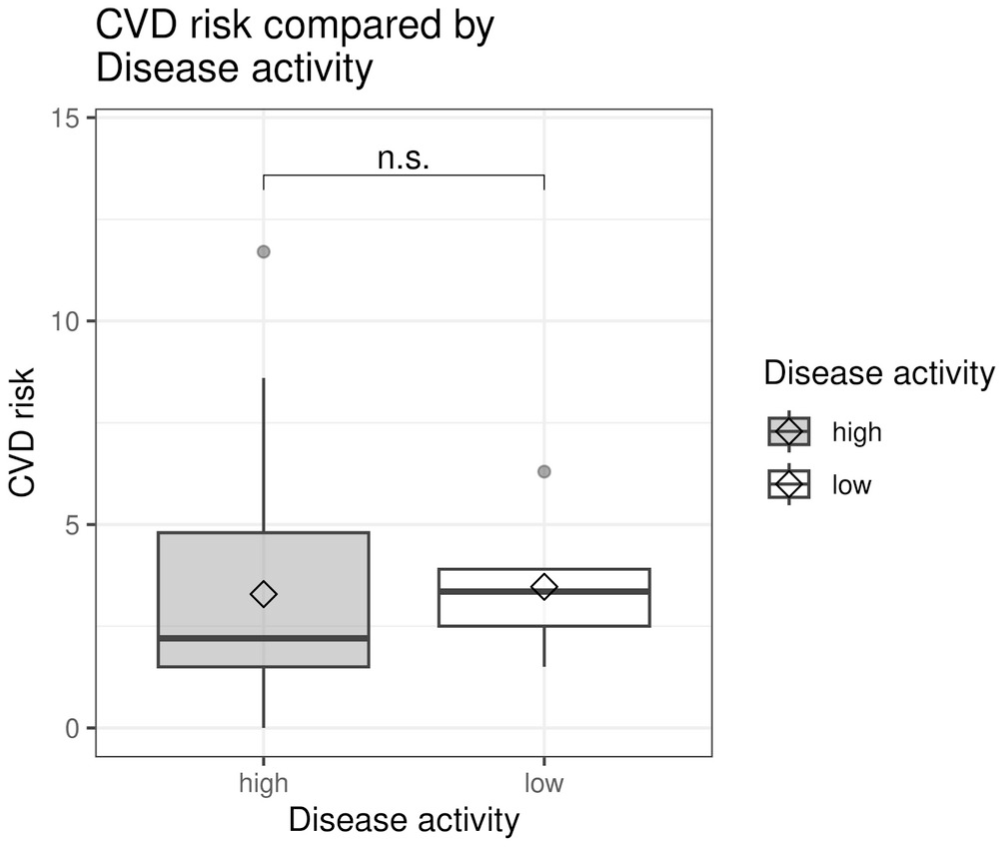
CVDs risk comparison for high vs. low disease activity in the Clinical cohort (n=28). For our Clinical cohort, there were no significant CVDs risk differences between participants with high disease activity vs. participants with low disease activity (two-sided test, p-value: 0.8513); suggesting that the disease activity effect in regression model-2 (see Table 3) is just a marginal effect.

### 3.2 Predictive model for CVDs risk components

Using the LupusRGMX data set (n=2914 participants) as our Registry cohort, we built predictive Bayesian networks for CVDs risk components. As a first step, two Random Forest models, and the Boruta package algorithm were implemented and combined to identify potential predictive variables to be used in the Bayesian network implementation. Table 4, shows the concurrent and most important potential variables.

**Table 4.** Selected variables with potential effect on nephritis and hypertension.

| ***Potential Effect on Nephritis*** | ***Potential Effect on Hypertension*** |
| --- | --- |
| Corticoids consumption | Deep vein thrombosis |
| Mycophenolic acid | Heart attack |
| Years with lupus | Peripheral arterial disease |
| Last ANA test | Diabetes |
| Methotrexate | Angina pectoris |
| Age | Corticoids consumption |
| Health provider | Comorbidities |
| Last anti-DNA, antiphospholipid, Smith, RNP, Ro, C3, or C4 test | Pulmonary thromboembolism |
| Socioeconomic level | Heart failure |
| Comorbidities | Socioeconomic level |
| Health satisfaction | State depression |
| Cyclophosphamide | Trait depression |
| Prednisolone | Coronary artery disease |
| Disease activity (measured by SLEDAI) | Quality of life perception |
| Quality of life perception | Insomnia |
| Occupation | Venous insufficiency |
| Diagnosis delay | Born prematurely |
| Vitiligo | Hashimoto's thyroiditis |
| Quality of life score | Gastroesophageal reflux disease |
| Chronic renal disease | Sleep apnea |
| Trait anxiety | Presbyopia |
| State depression | Pneumonia |
| Trait depression | Chronic renal disease |
| Hernias | Sjögren's syndrome |
| Pancreatitis | Gout disease |
| Polycystic ovary syndrome | Farsightedness disease |
| Anti dsDNA antibodies | Prednisolone |
| Anti-Sm anti-RNP anti-Ro antibodies | Gestational hypertension |
| Urolithiasis | Allergic rhinitis |
| Damage accrual (according to SLICC) | Trigeminal neuralgia |
| Venous insufficiency | Obsessive-compulsive disorder |
| Cannabis addiction | Somnambulism |
| Uterine myomatosis | Epilepsy |
| Inflammatory intestinal disease | Retinal detachment |
| Cholelithiasis | Hypertriglyceridemia |
| Presbyopia | Irritable bowel syndrome |
| Macular degeneration | Stroke |
| Azathioprine | Acute pancreatitis |
| Family with lupus | Hypercholesterolemia |
| aPL auto antibodies | Dyslexia |
| Psoriasis | Gestational diabetes |
| Myopia | Dry eye syndrome |

These variables (see Table 4) were used as inputs in the structure learning algorithms (i.e., GS, and ARACNE algorithms) for both Bayesian networks. These network structures were presented to rheumatologists and nephrologists for validation, including their expert knowledge in the network..

The Bayesian network shows how CRFs, SLE-associated factors, and psychosocial and sociodemographic factors contribute to nephritis in Mexican people with SLE (Figure 3). In particular, we observed: years with lupus, aPL autoantibodies, SLE diagnosis delay, C3 and C4 complement factors, damage accrual (according to SLICC), disease activity (measured by SLEDAI), anti-dsDNA antibodies, anti-Sm, anti-RNP, anti-Ro antibodies, relatives with SLE, methotrexate consumption, and mycophenolic acid consumption. Among psychological factors, we observed: trait anxiety, trait depression, state depression, and state anxiety. Regarding sociodemographic factors, we identified the following: age, socioeconomic level, health satisfaction, health provider, and quality of life. Other factors included those related to affected vision, as well as urinary or reproductive diseases. This network model performance was verified on the validation dataset (30% of the lupus nephritis dataset), giving an accuracy of 0.72, a precision of 0.67, a recall of 0.90, and a F1 score of 0.76.

**Figure 3.**
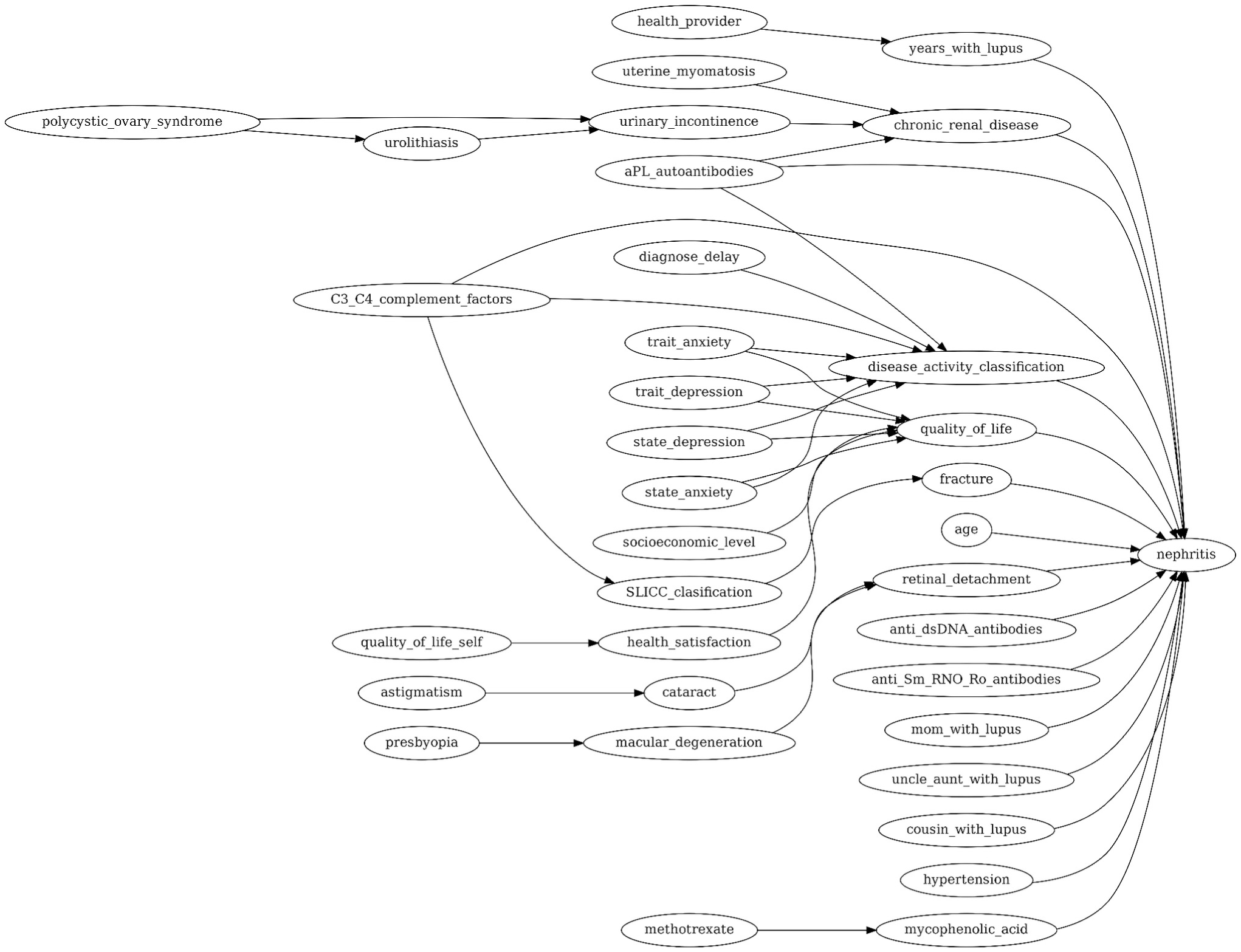
Nephritis network. Proposed Bayesian network of CRFs, SLE-associated factors, and psychosocial and sociodemographic factors, contributing to nephritis in Mexican people with SLE. Network performance was verified into the validation dataset (30% of the nephritis dataset), giving an accuracy of 0.72, a precision of 0.67, a recall of 0.90, and a F1 score of 0.76. The proposed network correctly identified 90% of the actual nephritis cases; however, it yields 37% of false-positive nephritis cases. Arrows depict the direction of such causality.

Our Hypertension Bayesian Model shows how CRFs, SLE-associated factors, including psychosocial and sociodemographic factors, contribute to this component to CVDs in people with SLE (Figure 4). In particular, among SLE-associated factors predicting nephritis, we identified the following as contributing factors: cyclophosphamide consumption, years with lupus, SLE diagnosis delay, damage accrual (according to SLICC) classification, Native American ancestry (self-reported), and disease activity (SLEDAI) classification. Regarding psychological factors, we observed depression, anxiety, and dyslexia. Among sociodemographic factors, we identified age, socioeconomic level, and quality of life. About traditional CRFs, we identified: tobacco use, alcohol use, obesity, overweight and eating behavior. Other factors included vision affections, pregnancy and reproductive health affections, as well as heart conditions. The network model performance was verified into the validation dataset (30% of the hypertension dataset), affording the following metrics: an accuracy of 0.82, a precision of 0.76, a recall of 0.94, and a F1 score of 0.84.

**Figure 4.**
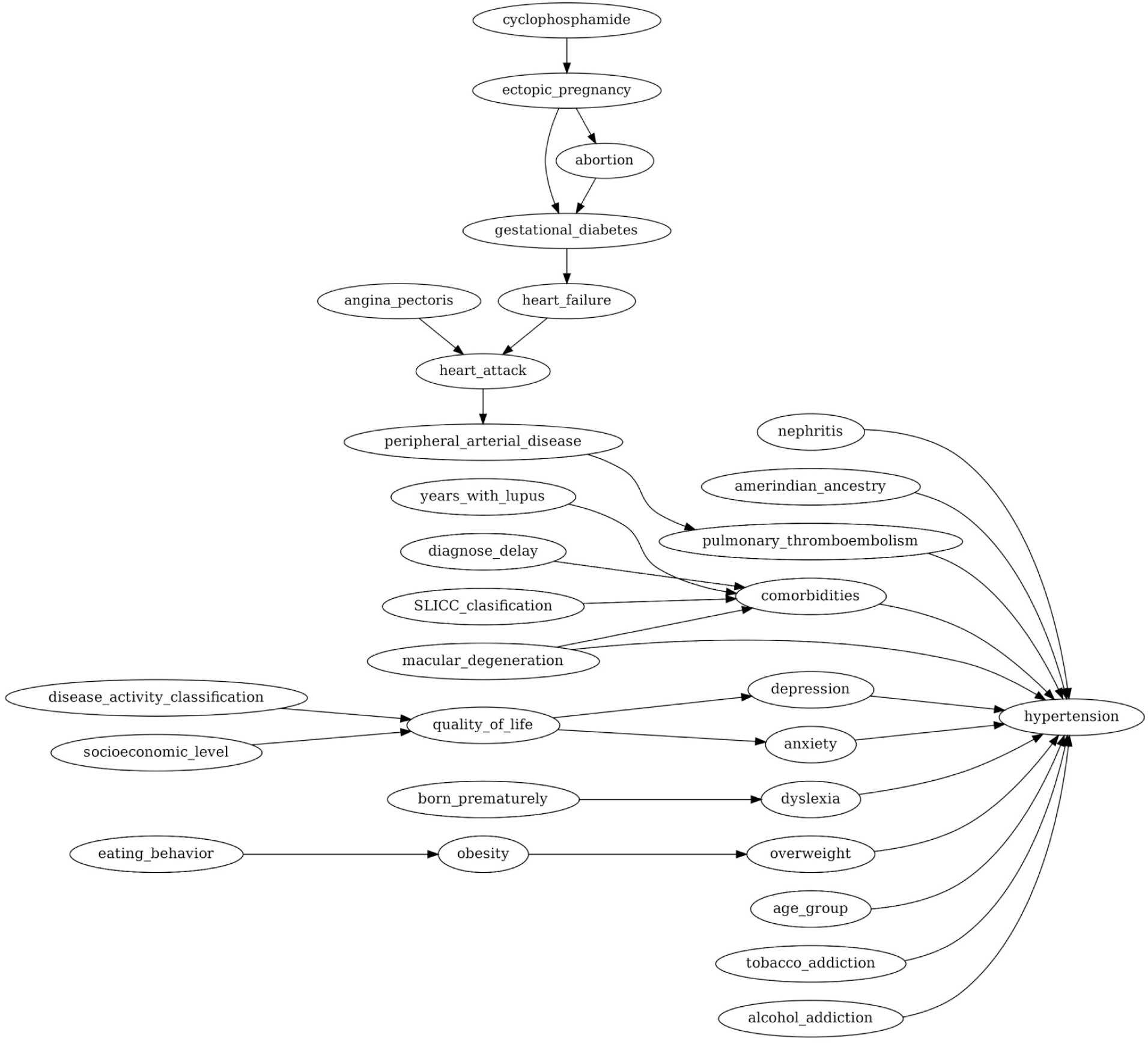
Hypertension network. Proposed Bayesian network of CRFs, SLE-associated factors, and psychosocial and sociodemographic factors, contributing to hypertension in Mexican people with SLE. Network performance was verified into the validation dataset (30% of the nephritis dataset), giving an accuracy of 0.82, a precision of 0.76, a recall of 0.94, and a F1 score of 0.84. The proposed network correctly identified 94% of the actual hypertension cases; however, it yields 24% of false positive hypertension cases. Arrows depict the direction of such causality.

Taking all results as a whole, we identified that classical CRFs are unable to fully account for the increase in cardiovascular risk observed in individuals with SLE, when such cardiovascular risk assessment is only based on the standard method of FRS.

## 4 Discussion

Although helpful, the assessment of cardiovascular risk through tools designed for the general population, such as the FRS, may overlook the contribution of risk factors inherent to SLE, underestimating the actual cardiovascular risk of people with SLE. The present study aimed to evaluate how CRFs and SLE-associated factors contribute to the development of CVDs in people with SLE. Through the assessment of the cardiovascular risk using the FRS as a standard method, we identify that classical CRFs cannot fully explain the increase in cardiovascular risk of people with SLE. By integrating the SLE-associated factors, such as nephritis diagnosis, years with SLE, diagnosis delay, treatment, and disease activity along with psychosocial and sociodemographic factors such as socioeconomic status and quality of life, we successfully integrated a predictive model to estimate the risk for the development of hypertension and nephritis in people with SLE.

In our Clinical cohort of 28 Mexican women with SLE, we evaluated the prevalence of classical CRFs and the FRS. We observed that, the majority of our volunteers presented mild and high risk (64.3% and 25%, respectively), results that contrast with a previous study conducted in the Mexican population, which reported 66.7% of their volunteers with low risk (40). Regarding the lipid profile, we observed that, against the expected, hypercholesterolemia (3.6%), hypertriglyceridemia (10.7%) and high levels of LDL-C (7.1%), were less prevalent in our cohort than in another Mexican population with SLE; in which, hypercholesterolemia (49%), hypertriglyceridemia (35.3%), and high levels of LDL-C (19.6%) were more frequent (40). The lower frequency of dyslipidemias among our clinical cohort in contrast with other Mexican cohorts, could be associated with different SLE-associated factors, including the use of treatments such as hydroxychloroquine and corticoids, the disease activity and accrual damage; which has not been fully explored in previous studies involving the Mexican population (41–43). For instance, in our clinical cohort, the majority (71%) were receiving antimalarial therapy, which has been associated with normal levels of lipid profile (42). When asked, twelve of the volunteers (42.8%) reported previously experiencing a cardiovascular event, being venous insufficiency (28.6%), and a hypertension diagnosis (25%), the most frequent.

Furthermore, in our Clinical cohort we ran two regression models; the regression model-1 considered traditional CRFs to explain CVDs risk, whereas the regression model-2 considered SLE-associated factors as explanatory variables of CVDs risk. The traditional CRFs model presented inconsistencies, for instance, tobacco use and high total cholesterol level showed a protective effect on CVDs risk, which is contra-intuitive. Since the protective effect of tobacco use was rejected by an additional permutation test, and our sample could erroneously bias the protective effect of high total cholesterol, we interpreted that traditional CRFs could exhibit a smaller effect on the CVD risk than SLE-associated factors. In other words, our analysis suggests that the impact of SLE-associated factors has a more prominent role in predicting CVDs risk than traditional factors, at least for the Mexican SLE population.

Even though both models were significant, the model composed of SLE-associated factors (regression model-2) better explains the CVDs risk, with an R-squared of 0.7955, in contrast, the model consisting of traditional CRFs, only got an R-squared of 0.5173. In other words, SLE-associated factors are better predictors of CVDs risk in the Mexican population. Specifically, years living with lupus, socioeconomic level, disease activity classification (measured by SLEDAI), age at diagnosis, corticoids and antimalarials consumption, are the SLE-associated factors that influence the CVDs risk the most. Being the last two, the factors with the most significant effects on CVDs risk.

It is worth mentioning that these last two factors, antimalarials and corticoids consumption, exhibited a contrasting effect on CVDs risk. Antimalarial consumption showed a protective effect, reducing Framingham Risk Score by 2.0 points, whereas corticoid consumption showed a risk effect, increasing Framingham Risk Score by 1.6 points, which is consistent with the literature (44,45). Moreover, disease activity has a marginal protective effect, but a permutation test did not support this effect.

Hence, regarding our Registry Cohort (n = 2914), when we integrate traditional CRFs, SLE-associated factors, psychological factors, and sociodemographic factors in a Bayesian Network, we can make better predictions for a major risk marker for CVDs, i.e., hypertension in lupus. Furthermore, bringing all these factors together, we can also successfully predict nephritis, which in turn is closely linked to hypertension in SLE.

When analyzing the Nephritis Bayesian Network (Figure 3) we identified several SLE-associated factors that contribute to nephritis prediction, including the use of mycophenolic acid, which was expected, as it is considered among the first-line drugs for lupus nephritis treatment (46–48). The presence of antibodies (anti-dsDNA, anti-Sm, anti-RNP, anti-RO) has also been associated with lupus nephritis (47,48). Specifically, anti-dsDNA antibody has been recognized to contribute to nephritis development and elevation of its levels has been linked to SLE flares, which can be represented in our model as disease activity (48). Other contributing factors were age and years with a lupus diagnosis. Previous studies have reported that nephritis is more likely to develop among the first years of diagnosis, which are mainly between 20 and 40 years, and it can evolve to chronic renal disease (49).

Regarding the Hypertension Bayesian Network (Figure 4) we identified traditional CRFs such as use of tobacco, eating behavior, obesity, and overweight. Interestingly, we observed that disease activity and socioeconomic level contribute to quality of life perception, which can reflect on the development of psychosocial traits such as depression and anxiety, and contribute to hypertension in our cohort. We have previously proposed that, in people with SLE, physical and socioeconomic burdens are associated with the capability of retaining employment and access to health services, which impacts on their quality of life perception and mental health (22). Previous studies have reported the association of depression and anxiety with CVDs in the general population; in SLE it has been proposed that it is mediated by inflammatory markers and influenced by lifestyle and some traditional CRFs such as overweight (50,51). Regarding lupus-associated factors, we found that years with lupus, diagnosis delay and damage accrual (according to SLICC) contribute to the development of comorbidities, including hypertension. In line with this, delay in the diagnosis has been associated with chronic inflammation, which can lead to organ damage and the development of conditions such as lupus nephritis and CVDs (52).

An interesting result to highlight is the close relationship between lupus nephritis and hypertension in people with SLE. We identified in both Bayesian Networks that these two factors interact with each other and contribute to their development in people with SLE (Figure 3 and Figure 4). Previous studies have identified that, in people with SLE, the incidence of hypertension increases along with accumulated kidney damage (12). It is worth noting that some SLE-associated factors, as well as psychological and demographic factors, are present as causal factors in both networks. These factors are: disease activity (SLEDAI), damage accrual (according to SLICC), years with lupus, diagnosis delay, anxiety, depression, quality of life, and socioeconomic level. Further studies should deepen the relationship between lupus nephritis and the development of hypertension in people with SLE, as well as identify the contribution of lupus-associated factors.

Concerning performance metrics of the models, the nephritis model correctly predicted 72% of all cases, i.e., people with and without nephritis; whereas, the hypertension model correctly predicted 82% of all cases of people with and without hypertension. Regarding precision, in the nephritis model, 67% of the cases predicted as nephritis were actually correct; whereas, in the hypertension model, 76% of the cases predicted as hypertensive were actually correct. In both models, we had a relatively high rate of false positives, 37% of false positive nephritis cases, and 24% of false positive hypertension cases. On the other hand, recall is relatively good for both models, the nephritis network correctly identified 90% of the actual nephritis cases, whereas the hypertension network correctly identified 94% of the actual hypertension cases. Finally, the F1-score metric balances precision and recall, giving a single measure of overall performance, the greatest F1-score means that the network model is better balanced. Then, both network models got an acceptable balance, 0.76 F1-score for the nephritis network, and 0.84 F1-score for the hypertension network.

Speaking of limitations, this study faced three relevant limitations. First, we performed cross-sectional analyses. Second, in our Clinical cohort (n=28), some variables, such as hypercholesterolemia, were imbalanced, which could limit the models’ capacity to identify potential effects. Third, in the models of our Registry cohort (n=2914), we had a relatively high rate of false-positive cases classified as nephritis or hypertension and perhaps attributed to the lack of additional essential biomarkers of nephritis and hypertension. In this line, future approaches would benefit from including additional molecular biomarkers. For instance, transcriptional profiles, specifically, neutrophil signatures, have been reported to be associated with progression to nephritis (53). Other transcriptomics and microarray approaches have shown that type I interferon (IFN) pathways are up-regulated in SLE patients, which is strongly connected to vascular disease (54) and pulmonary arterial hypertension (55). These molecular biomarkers should be included longitudinally in future studies.

## 5 Conclusion

People with SLE face a higher risk of developing CVDs. While traditional cardiovascular risk factors are more prevalent in people with SLE, these alone do not fully account for the increased CVDs risk observed in this population. By analyzing two cohorts, this study integrated data that allowed to identify that, beyond conventional CRFs, lupus-associated factors -as well as psychological and sociodemographic variables-play a key role in estimating cardiovascular risk in people with autoimmune diseases like SLE. Using data from the LupusRGMX cohort, two Bayesian network models were designed and evaluated, showing that lupus-associated factors significantly contribute to the development of hypertension and nephritis, two leading causes of morbidity and mortality in SLE. Key variables such as treatment, diagnosis delay, years with the disease, damage accrual, disease activity, antibody levels, self-reported ancestry, and family history of lupus should be integrated into CVDs risk assessment models for individuals with lupus. This approach will help identify the underlying factors that contribute to the development of severe, potentially life-threatening complications in people with SLE. This will facilitate a more preventive strategy for disease management, improving the long-term outcomes and the overall quality of life for people living with SLE.

## 7 Author Contributions

ALH-L: Conceptualization, Data curation, Investigation, Formal analysis, Writing – original draft, Writing – review & editing, SFH-R: Conceptualization, Data curation, Investigation, Resources, Writing – original draft, ASDAZ: Conceptualization, Data curation, Investigation, Resources, Writing – original draft, GS-P: Data curation, Formal analysis, Validation, Visualization, Writing – original draft, AP-A: Conceptualization, Resources, Writing – review & editing, LT-N: Conceptualization, Resources, Writing – review & editing, ET-V: Conceptualization, Resources, Writing – review & editing, DGG-G: Investigation, Resources, Writing – review & editing, AJR-M: Investigation, Resources, Writing – review & editing, JS-D: Validation, Writing – review & editing, GMV-M: Validation, Writing – review & editing, DA-R: Conceptualization, Validation, Writing – review & editing, AM-R: Supervision, Funding acquisition, Writing – review & editing, DM: Conceptualization, Data curation, Formal analysis, Methodology, Software, Visualization, Writing – original draft, Writing – review & editing.

## 9 Acknowledgments

This work received support from Luis Aguilar, Alejandro León, and Jair García of the Laboratorio Nacional de Visualización Científica Avanzada. We also thank Carina Uribe Díaz, Alejandra Castillo Carbajal and Christian Molina Aguilar for their technical support. Authors would like to express their special acknowledgment to Fundación Proayuda Lupus Morelos A.C, Lupus MX, El despertar de la Mariposa, and Centro de Estudios Transdisciplinarios Athié-Calleja por los Derechos de las Personas con Lupus A.C., for their invaluable support.

## 11 Data Availability Statement

All data is available upon request through the MexOMICS Consortium (https://redcap.link/nqsxtj8n). The complete analysis code is avialibale through GitHub https://github.com/NeuroGenomicsMX/Cardiovascular_Risk_Factors_Lupus_Mex.

